# The clinical efficacy of faecal microbiota transplantation: An umbrella review of randomized controlled trials

**DOI:** 10.1101/2021.11.18.21266556

**Authors:** Nanyang Liu, Tingting Zhang, Jiahui Sun, Jianhua Fu, Hao Li

**Author notes:** Correspondence Hao Li, PhD, Wangjing Hospital, China Academy of Chinese Medical Sciences, Beijing, China, Jianhua Fu, PhD, Xiyuan Hospital, China Academy of Chinese Medical Sciences, Beijing, China. **Guarantor of the article:** Hao Li PhD. **Specific author contributions:** Nanyang Liu, and Jianhua Fu conducted the literature search, study selection and data extraction and analysis. Tingting Zhang, Jiahui Sun, Jianhua Fu and Hao Li contributed to the data interpretation, writing, and editing of the manuscript. All authors have read and agreed to the published version of the manuscript.

## Abstract

**INTRODUCTION:** Faecal microbiota transplantation (FMT) therapeutic strategy has been associated with positive outcomes in multiple diseases. We conducted an umbrella review of the meta-analysis to summarize the available evidence and assess its credibility.

**METHODS:** We evaluated a meta-analysis of randomized controlled trials that investigated the efficacy and safety of FMT for any condition. We used the random-effects model to obtain estimates and corresponding 95% confidence intervals, heterogeneity estimates, and small-study effects. We used AMSTAR 2 to assess methodological quality and GRADE tools to grade the evidence.

**RESULTS:** Seven meta-analyses with a total of 33 outcomes were included in the current umbrella review to evaluate the efficacy and safety of FMT. Overall, there is a moderate certainty of evidence supporting the associations of the use of FMT with better clinical remission in patients with Clostridium difficile infection (RR = 1.74; 95% CI: 1.37-2.22) and inflammatory bowel disease (RR = 1.70; 95% CI: 1.12-2.56). A very low certainty evidence supports the use of FMT to treat antibiotic-resistant bacteria (RR = 5.67; 95% CI: 2.20-14.63) and functional constipation (RR = 1.35; 95% CI: 1.14-1.60) but does not support irritable bowel syndrome (RR = 1.08; 95% CI: 0.65-1.77).

**DISCUSSION:** The umbrella review of the current meta-analysis demonstrates that FMT intervention is associated with positive outcomes for several diseases. However, the credibility of the evidence is not high. Further high-quality randomized controlled trials should be carried out to improve the strength and credibility of the evidence base.

## INTRODUCTION

Accumulating evidence emphasizes the potential contribution of commensal gut microbiota in human health and various diseases like inflammatory bowel disease (IBD)^1, 2^, irritable bowel syndrome (IBS)^3, 4^, and gastrointestinal cancer^5, 6^. It is also important in nongastrointestinal diseases, such as cardiovascular^7^, metabolic^8^, neurological^9^, and psychiatric diseases^10^. In the past two decades, microbiology has developed at an alarming rate and has revealed the various ways in which these tiny organisms affect our health. Advances in sequencing technology coupled with updates to the microbiome information pipeline have made microbiome analysis cheaper and more complex. On this basis, the interaction mechanism between commensal microbiota and these diseases has been gradually revealed. In this context, recent positive evidence supports the use of antibiotics, prebiotics, probiotics, or faecal microbiota transplantation (FMT) to prevent or treat microbiota-associated diseases^11^, and achieved some impressive results.

FMT is an emerging therapeutic method that has become a research hotspot in biomedicine and clinical medicine^12^. The process includes transplanting functional microbiota from healthy individuals into the intestinal tract with pathological microbiota to improve dysbiosis, thereby treating intestinal and extraintestinal diseases. FMT was originally used to treat pseudomembranous colitis caused by Clostridium difficile infection (CDI). Recently, it has been approved as the standard treatment therapy for recurrent CDI by official guidelines due to its remarkable curative effect^13^. Emerging evidence links gut microbiota disorders with the pathology of numerous diseases^14^, prompting researchers to continue to expand the scope of this strategy. According to the latest data from clinicaltrials.gov, nearly 400 trials involving nearly 100 diseases or conditions have been completed or are in progress, most of which were conducted in the past five years. In addition to well-known gastrointestinal disorders, cardiovascular and neurologic diseases are also attracting attention **(Figure 1)**.

**Figure 1.**
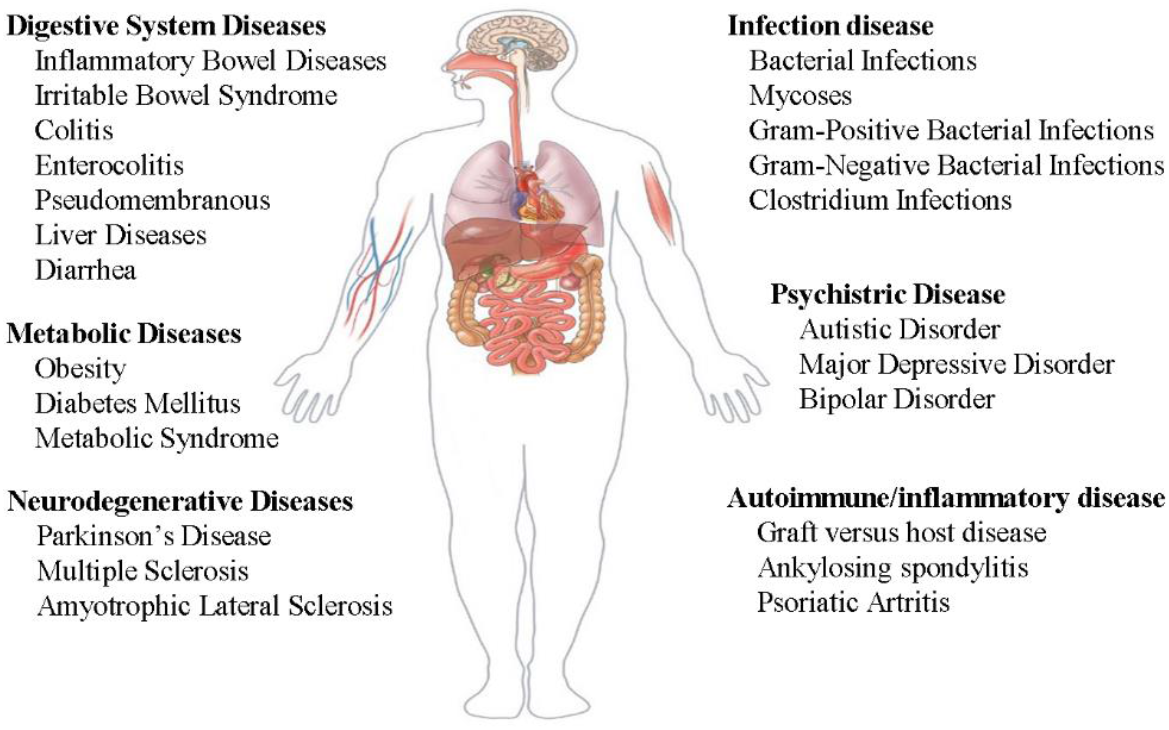
Ongoing clinical trials on faecal microbial transplantation. The data comes from www.clinicaltrial.gov.

In recent years, evidence from randomized clinical trials (RCTs) on the efficacy and acceptability of FMT has been obtained via both meta-analyses and network meta-analyses^15-18^. However, no research has attempted to quantify the credibility of these findings to date. The umbrella review aims to summarize evidence from multiple meta-analyses on the same topic and evaluate sample size, strength of association, and risk of bias to rank the evidence^19-21^. In this context, we conducted an umbrella review of existing meta-analyses to quantify the strength of the association between FMT and any conditions. We summarized the findings of the cross-meta-analyses, assessed methodological quality, and investigated potential biases to determine which outcomes are supported by reliable evidence.

## METHODS

### Search Strategy

The systematic literature search was conducted in PubMed, Embase, and Cochrane Library from database inception to August 1, 2021, to identify meta-analyses of RCTs investigating the intervention effect of FMT on any disease. The search strategy used a combination of the following terms: faecal microbiota transplantation (e.g., intestinal microbiota transfer, faecal transplantation, donor faeces infusion) and meta-analysis (e.g., systematic review, meta-analysis, review). No restrictions or filters were applied for the search process. We also manually searched the cited references of the retrieved articles and reviews. Two authors independently conducted literature search. Any disagreements were resolved by consultation with a third author. The detailed search strategy is provided in **Supplementary Table 1**.

### Selection Criteria

Systematic reviews with meta-analyses of RCTs were included, regardless of the frequency, dosage, and transmission forms of faeces. For multiple meta-analyses of the same result, we selected only one meta-analysis for each result to avoid including duplicate studies^22^. In this case, we included the largest number of primary studies. If more than one published meta-analysis included the same number of studies, then the one with the largest number of patients was selected. If more than one published meta-analysis meets these two criteria, we selected the one with more available information (e.g., dose-response meta-analysis)^22^. The competitive meta-analysis was screened to identify any additional trials not included in the selected meta-analysis. When qualified studies contained multiple types of results, we only extracted the pooled effect estimates of RCTs^23, 24^. Additionally, we also searched the latest primary study to avoid missing updated data.

Studies were excluded if they were network meta-analyses, if they were systematic reviews without meta-analyses, if the full text of the meta-analysis was not available, or if the meta-analysis lacked data for summary estimates.

### Data Extraction

Two authors independently extracted data, and disagreements were resolved by consensus. From each meta-analysis, we extracted the first author, journal name, publication year, study design, type of comparison, interesting outcomes, and the number of included studies. We also extracted relative risk (RR) estimates, odds ratio (OR), 95% confidence intervals (CI) and corresponding *P* values, the number of participants and events, follow-up time, meta-analysis models used (fixed effects or random effects), and information on heterogeneity, small-study effects, funding, and conflict of interest. We also extracted any recorded subgroup analysis estimates. For each primary study that was additionally retrieved, the above data were also extracted to update the meta-analysis.

### Assessment of methodological quality

We used AMSTAR 2 (A Measurement Tool to Assess Systematic Reviews 2), a strict, validated, and reliable measurement tool, to assess the methodological quality of each meta-analysis^25^. It consists of 16 items, of which 7 are key items, including quality ratings for meta-analysis of search, reporting, analysis, and transparency^26^. According to the weakness of the key items, the methodological quality was assessed on 4 grades: high, moderate, low, or critically low^26^ **(Methods in Supplementary Table 2)**.

### Evaluation of quality of evidence

We used the GRADE (Grading of Recommendations, Assessment, Development and Evaluation) assessment to evaluate the credibility of the evidence provided by each association in the meta-analysis^27, 28^. Evidence from the meta-analysis of randomized controlled trials was evaluated based on the significance of the pooled effect, using a *p value* of <0.05 as statistical significance. The unreported *P value* was calculated from the 95% confidence interval of the collective effect estimate by using standard methods.

### Statistical Analysis

The effect sizes of individual studies included in each meta-analysis were extracted when the reported data were sufficiently detailed. After removing duplicate trials, we updated each meta-analysis by combining the latest published data using the DerSimonian and Laird random-effects models, which considers variance between and within studies^19^. The interesting dichotomous variables are clinical remission and total effective rate, which best reflect the clinical efficacy of FMT. Meta-analysis with multiple continuous variables is displayed in one forest plot. We did not review the primary study included in each meta-analysis. We used the logarithmic scale to calculate the summary estimate and then performed exponential analysis to return the result to the original indicator. Heterogeneity between studies was assessed using *I*^2^ statistics. Values < 50% indicate acceptable heterogeneity, values > 50% suggest high heterogeneity, and values > 75% are indicative of high heterogeneity^19^. Egger’s regression asymmetry test was used to calculate an estimate of publication bias for any reanalysis that included at least 10 studies, which was considered indicative of small-study effects^26^. A *P value* < 0.1 was considered statistically significant by Egger’s test.

## RESULTS

### Search Results

The initial systematic search identified 244 records. After deleting duplicates, we reviewed the titles and abstracts of all retrieved articles, and finally, 87 (includes 53 meta-analyses) were determined. Considering the purpose of the present umbrella review, we selected those studies that included the largest number of RCTs. Ultimately, 7 meta-analyses met the eligibility criteria^29-35^. **Figure 2** shows the flowchart of the literature search. A list of excluded studies can be found in **Supplementary Table 3**.

**Figure 2.**
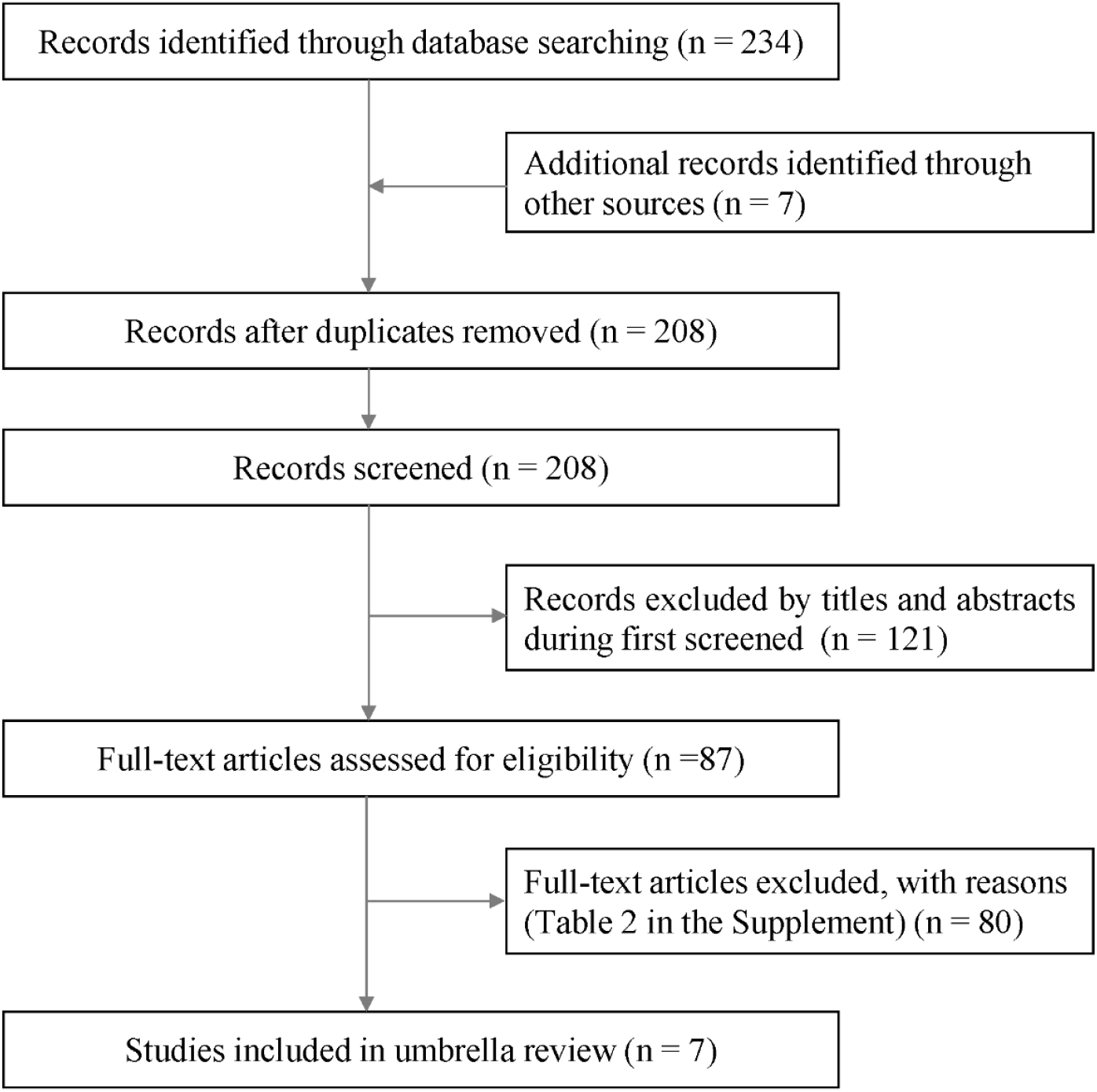
The flowchart of the literature search.

### Characteristics of meta-analyses

The populations considered were in 7 meta-analyses people with CDI^33^, IBD^31^, IBS^34^, ulcerative colitis (UC)^32^, functional constipation^30^, metabolic syndrome^35^, and antibiotic-resistant bacteria^29^. The meta-analyses were published between 2019 and 2021. The median number of primary studies per meta-analysis was 6 (interquartile range, 6-8), and the median number of cases was 355 (interquartile range, 147-471). The FMT group mostly used fecal microbiota from healthy donors, while the control group mainly used placebo. Thirty-three effect estimates were reported in 7 meta-analyses, 17 of which recorded significant *P* < 0.05 effects. The general characteristics of the included meta-analysis can be found in **Supplementary Table 4**.

### Meta-analyses of RCTs

In placebo-controlled analysis^29, 31, 34, 35^, FMT had statistical clinical significance in the improvement of IBD (RR=1.70; 95% CI: 1.12-2.56) and antibiotic-resistant bacteria (RR=5.67; 95% CI: 2.20-14.63). There is no evidence that FMT has a beneficial effect on IBS (RR = 1.08; 95% CI: 0.65-1.77) outcome, although three recently published RCTs were included in the analysis^36-38^ **(Figure 3)**. For metabolic syndrome, FMT contributed to an increase in high-density lipoprotein and a decrease in low-density lipoprotein but did not change the parameters of glycosylated hemoglobin, fasting blood glucose, body weight, body mass index, triglycerides, or total cholesterol **(Figure 4)**.

**Figure 3.**
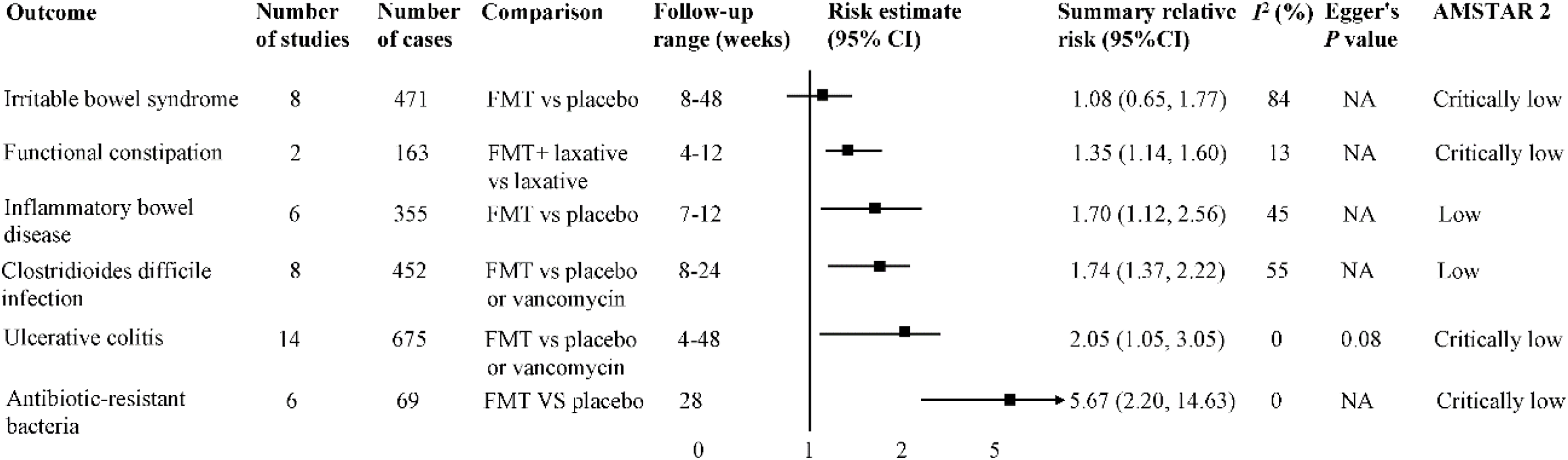
The association between FMT and multiple disease outcomes. Estimates are relative risks of reanalysis and effect models are random.

**Figure 4.**
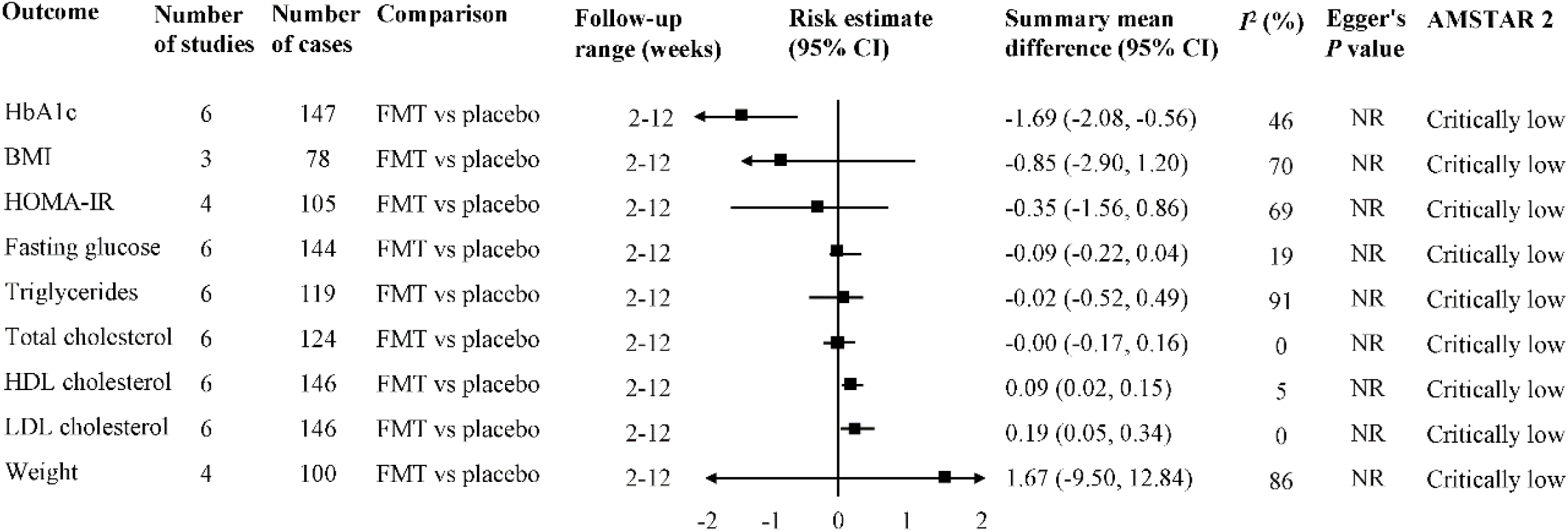
The association between FMT and metabolic syndrome. Estimates are relative risks of reanalysis and effect models are random.

In a meta-analysis of placebo- or vancomycin-controlled studies^32, 33^, the use of FMT was associated with better clinical remission of UC (RR=2.05; 95% CI: 1.05-3.05) and CDI (RR=1.74; 95% CI: 1.37-2.22). One recently published RCT was included in the analysis of CDI^39^ **(Figure 3)**.

The efficacy of FMT on functional constipation was limited to five RCTs in one meta-analysis^30^. In the control group administered laxatives, FMT combined with laxatives was more effective in relieving the symptoms of constipation (RR=1.35; 95% CI: 1.14-1.60) **(Figure 3)**.

### Methodological quality

The results of AMSTAR 2 for each meta-analysis are presented in **Supplementary Table 5**. Overall, the methodological quality assessment of 7 studies was determined to be critically low (5 studies, 71.4%) or low (2 studies, 28.6%). The most common critical flaws were the absence of a detailed literature exclusion list and funding sources and did not consider the risk of bias and heterogeneity when preparing conclusions and recommendations. However, due to the insufficient number of primary studies in the 6 meta-analyses (less than 10), publication bias was not evaluated, which may magnify the low methodological quality.

### Quality of Evidence

**Table 1** reports the GRADE approach to categorize the level of evidence for the significant outcomes of the included disease. Overall, moderate certainty of evidence indicated that the use of FMT was associated with a better CDI, UC, and IBD. A very low certainty of evidence supported the use of FMT in improving antibiotic-resistant bacteria and functional constipation. Moreover, very low-quality evidence indicated a lack of support for the use of FMT to treat IBS. The outcomes of metabolic syndrome were supported by different degrees of certainty, ranging from moderate to very low^35^.

**Table 1.**
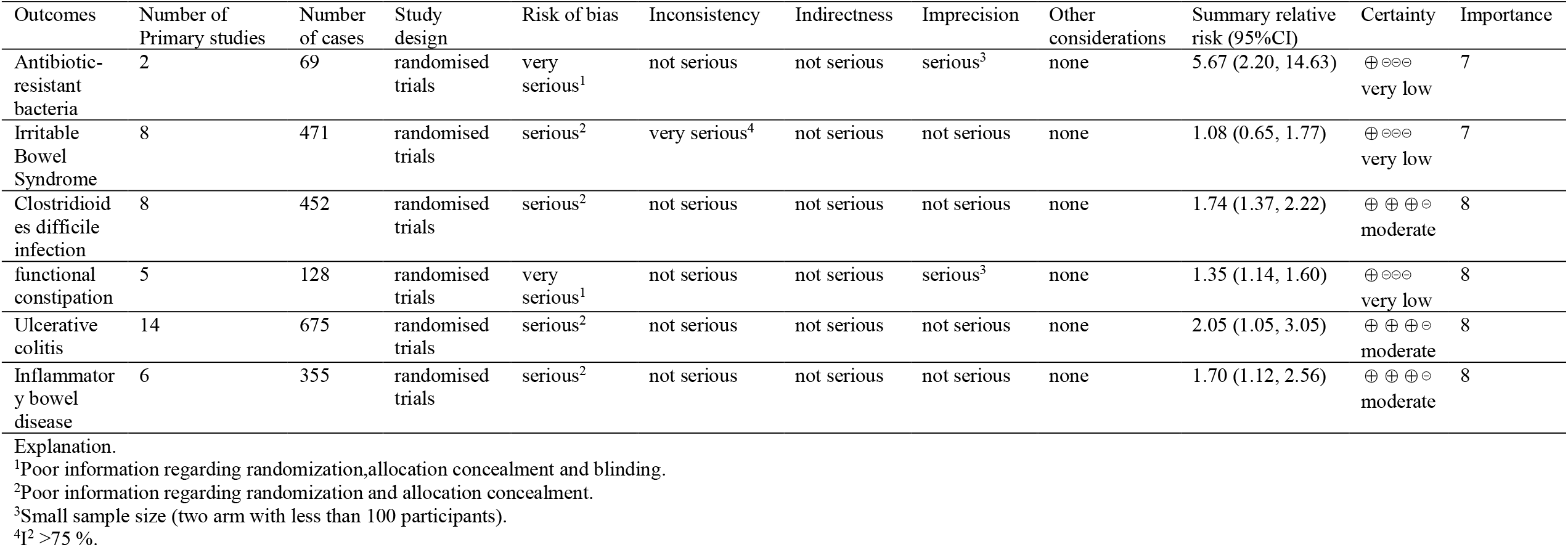
GRADE evidence for the included meta-analysis.

### Heterogeneity between primary studies

Significant heterogeneity was detected in the meta-analysis of IBS, while heterogeneity among other studies was acceptable. Most of the heterogeneity in the meta-analysis of functional constipation is acceptable.

### Publication bias and small study-effects

The publication bias and small study effects for each meta-analysis were evaluated by Egger tests. *P value*s < 0.10 are considered statistical evidence. Our results indicate the presence of small-study effects (potential publication bias) for UC. More than 10 primary studies were available for one meta-analysis, while the others were insufficient, and publication bias was not evaluated **(Figure 3 and 4)**.

## DISCUSSION

This review aims to provide a comprehensive overview of the efficacy and safety of FMT in gut dysbiosis-associated diseases by incorporating the evidence from the current meta-analysis. The present umbrella review, including 7 RCT meta-analyses, found that the FMT treatment strategy was significantly associated with multiple positive effects. Specifically, moderate evidence supports the use of FMT to treat CDI, UC, and IBD. Very-low-quality evidence supports the use of FMT to treat antibiotic-resistant bacteria and functional constipation but does not support the treatment of IBS. However, the current low-to-moderate evidence cannot determine the efficacy of FMT to treat metabolic syndrome. Of note, although positive results were obtained, the credibility was reduced due to the low quality of the evidence.

The current work supports and expands on a recent review that considered studies as of 2018^40^. This review highlights the positive effects of FMT on recurrent CDI, IBS, and IBD. Based on the evidence at the time, the author also suggested that patients with metabolic syndrome should be cautious when using FMT therapy, which is consistent with our point of view. However, our quantitative analysis does not support the use of FMT in patients with IBS. Inconsistent understanding allows us to continue to explore the potential impact of FMT on gut dysbiosis-associated diseases.

The safety assessment of FMT reached a satisfactory conclusion. Compared with placebo, adverse events in the FMT group were not statistically significant in the studies of Ianiro et al. (OR = 1.37, 95% CI 0.63–2.96) and Tang et al. (RR = 0.93 (95% CI 0.45– 1.92). Some common adverse events are mild and spontaneously relieved, like abdominal distension, diarrhea, nausea, abdominal pain, and fever. No serious adverse events were reported in the remaining 5 meta-analyses, indicating that FMT appears to be safe.

The gut microbiota refers to the bacteria, viruses, parasites, and fungi colonizing the intestinal tract^41^. The adult gut microbiota is composed of more than 2,000 bacteria to form a diversified, stable, resistant, and elastic microbial ecosystem that participates in host immunity, metabolism, and other biological functions^42, 43^. Dysbiosis is disturbances in the function and composition of the microbiota driven by environmental and host-related factors^12^. This process may be involved in the pathogenesis of many diseases, such as IBD, IBS^44^, multiple sclerosis^45, 46^, hepatic encephalopathy^47, 48^, cancer^49-51^ and metabolic syndrome^52^. Targeting the disturbed microbiota, which may be achieved by dietary interventions, probiotics, prebiotics, antibiotics, and FMT, might affect the progress of these conditions. Research related to FMT can obtain the most convincing evidence that gut microbiota plays a role in human diseases.^12^.

The application of stool therapy can be traced back to ancient Chinese medicine nearly 1700 years ago^53^. Eiseman et al. first reported FMT as an adjuvant treatment for patients with antibiotic-associated diarrhea, which opened the door to the modern era^54^. Subsequent reports confirmed that Clostridium difficile was the culprit responsible for post-antibiotic colitis (known today as pseudomembranous colitis)^55, 56^. Following these revelations, numerous trials indicated the clinical effect of FMT on pseudomembranous colitis caused by CDI and finally approved it as a standard treatment strategy by official guidelines^13^. With the continuous advancement of gut microbiota research, the underlying mechanisms of many conditions have been linked. Correcting the imbalanced gut microbiota is also becoming a potential alternative strategy. Recently, FMT treatment attempts have gradually expanded from the initial gastrointestinal disorder to other diseases, such as the nervous system and cardiovascular system. Additionally, the establishment of a stool bank makes FMT an easily available and useful option.

Although this novel approach seems safe and easy to implement, we should be cautious because the long-term effects are still unknown or unrecognized. Moreover, as an emerging medical therapeutic strategy, FMT is not yet a standardized treatment method. The protocols vary according to local procedures. Uniform standards not established on faecal formulation, transplantation method and frequency may be the reason for the inconsistent results.

This umbrella review used the AMSTAR 2 tool to assess the methodological quality of meta-analysis and identified several potential flaws, like inaccurate assessments of risk of bias and heterogeneity, a lack of funding sources, and literature exclusion lists. These flaws lead to the low quality of the evidence from primary studies, thereby affecting the overall quality (low or very low) of the meta-analysis. Insufficient information on randomization, allocation concealment and blinding are the main factors that downgrade the quality of evidence. This was followed by small sample size and significant heterogeneity. We did not find any convincing factors to upgrade the quality of evidence. Of note, future meta-analyses in this field should use AMSTAR 2 as an executive checklist to ensure high-quality evidence.

Our umbrella review had several strengths. First, we provide a systematic and comprehensive overview of all published meta-analysis evidence regarding the role of FMT in gut dysbiosis-associated diseases. Second, we assessed the methodological quality and quality of evidence using validated tools. In this case, although the quality of most meta-analyses was not high, some internal flaws were revealed, and future research directions could therefore be identified. Third, we additionally searched for the latest trials published after the previous meta-analysis to avoid missing key information. Overall, our review, including recent meta-analysis and previously unreported findings, may represent significant evidence for further research and policy development.

Potential limitations should be considered when interpreting the results of our work. First, we used pre-established tools to assess the quality of meta-analysis, which relies on complete data in the primary study. Although the two authors conducted the assessment back-to-back, subjectivity was inevitable. Second, since the focus of our umbrella review is to provide broad-based evidence from existing meta-analyses, subgroup analysis (e.g., by fecal formulation, transplantation method, and frequency) and sensitivity analysis (e.g., to exclude studies with a high risk of bias) were not performed. For instance, fresh fecal bacteria are more effective than cold storage, and multifrequency transplantation lasts longer than single transplantation. Third, only one meta-analysis applied the Egger’s test (small study effect). Most meta-analyses included 5 to 10 primary studies, even one included fewer than 5 studies. Some data may be missing due to the low number of primary studies. Fourth, we used DerSimonian and Laird’s random-effects method to calculate the aggregate hazard ratio and the corresponding 95% CI to ensure comparability with the previous meta-analysis. However, future meta-analyses should use the Hartung-Knapp method, which can better reflect the uncertainty of the differences between studies, expressed with a wider confidence interval.

## CONCLUSION

This umbrella review summarized the current evidence on the safety and efficacy of FMT in microbiota-associated disease and found several positive associations. Our work highlights the significance of the use of FMT by public health authorities in some diseases associated with gut microbiota disorders. However, low-to-moderate quality of evidence makes it necessary to be cautious when adopting recommendations. Continued research into the therapeutic effects of FMT is important since the gut microbiota interacts with the pathophysiology of many diseases. In addition to large-scale RCTs, well-designed long-term follow-up protocols must also be considered to evaluate longer-term efficacy and safety.

## Data Availability

All data produced in the present study are available upon reasonable request to the authors

