## Supplementary material for "The clinical efficacy of faecal microbiota transplantation: An umbrella review of randomized controlled trials": NA

| Table 1. Detailed search query |
|---|
| **Database:** PubMed<br>**Date limits:**<br>Fecal Microbiota Transplantation[MeSH] Fecal Microbiota Transplantations [Title/Abstract] OR Microbiota Transplantation, Fecal [Title/Abstract] OR Microbiota Transplantations, Fecal [Title/Abstract] OR Transplantation, Fecal Microbiota [Title/Abstract] OR Transplantations, Fecal Microbiota [Title/Abstract] Intestinal Microbiota Transfer [Title/Abstract] OR Intestinal Microbiota Transfers [Title/Abstract] OR Microbiota Transfer, Intestinal [Title/Abstract] OR Microbiota Transfers, Intestinal [Title/Abstract] OR Transfer, Intestinal Microbiota [Title/Abstract] OR Transfers, Intestinal Microbiota [Title/Abstract] OR Fecal Transplantation [Title/Abstract] OR Fecal Transplantations [Title/Abstract] OR Transplantation, Fecal [Title/Abstract] OR Transplantations, Fecal [Title/Abstract] OR Fecal Transplant [Title/Abstract] OR Fecal Transplants [Title/Abstract] OR Transplant, Fecal [Title/Abstract] OR Transplants, Fecal [Title/Abstract] OR Donor Feces Infusion [Title/Abstract] OR Donor Feces Infusions [Title/Abstract] OR Feces Infusion, Donor [Title/Abstract] OR Feces Infusions, Donor [Title/Abstract] OR Infusion, Donor Feces [Title/Abstract] OR Infusions, Donor Feces [Title/Abstract] AND Systematic review[Title/Abstract]) OR (meta-analysis[Title/Abstract] |
| **Database:** Embase<br>**Date limits:**<br>‘bacteriotherapy (feces)’:ab,ti OR ‘faecal bacteriotherapy’:ab,ti OR ‘faecal enema’:ab,ti OR ‘faecal infusion’:ab,ti OR ‘faecal matter transplant’:ab,ti OR ‘faecal microbial transplant’:ab,ti OR ‘faecal microbial transplantation’:ab,ti OR ‘faecal microbiome transplant’:ab,ti OR ‘faecal microbiome transplantation’:ab,ti OR ‘faecal microbiota transplant’:ab,ti OR ‘faecal microbiota transplantation’:ab,ti OR ‘faecal transplant’:ab,ti OR ‘faecal transplantation’:ab,ti OR ‘fecal bacterial transplant’:ab,ti OR ‘fecal bacterial transplantation’:ab,ti OR ‘fecal bacteriotherapy’:ab,ti OR ‘fecal enema’:ab,ti OR ‘fecal infusion’:ab,ti OR ‘fecal instillation’:ab,ti OR ‘fecal matter transplant’:ab,ti OR ‘fecal matter transplantation’:ab,ti OR ‘fecal microbe transplant’:ab,ti OR ‘fecal microbial transplant’:ab,ti OR ‘fecal microbial transplantation’:ab,ti OR ‘fecal microbiome transplant’:ab,ti OR ‘fecal microbiome transplantation’:ab,ti OR ‘fecal microbiota transplant’:ab,ti OR ‘fecal microbiotal transplant’:ab,ti OR ‘fecal microflora transplantation’:ab,ti OR ‘fecal transfusion’:ab,ti OR ‘fecal transplant’:ab,ti OR ‘fecal transplantation’:ab,ti OR ‘feces bacteriotherapy’:ab,ti OR ‘feces microbe transplantation’:ab,ti OR ‘feces microbiota transplantation’:ab,ti OR ‘feces microflora transplant’:ab,ti OR ‘feces microflora transplantation’:ab,ti OR ‘FMT (fecal microbiota transplantation)’:ab,ti OR ‘gut microbial transplant’:ab,ti OR ‘gut microbial transplantation’:ab,ti OR ‘gut microbiome transplant’:ab,ti OR ‘gut microbiome transplantation’:ab,ti OR ‘gut microbiota transplant’:ab,ti OR ‘gut microbiota transplantation’:ab,ti OR ‘gut microflora transplantation’:ab,ti OR ‘IMT (intestinal microbiota transplantation)’:ab,ti OR ‘intestinal microbe transplantation’:ab,ti OR ‘intestinal microbiota transplant’:ab,ti OR ‘intestinal microbiota transplantation’:ab,ti OR ‘intestinal microflora transplantation’:ab,ti OR ‘rectal bacteriotherapy’:ab,ti OR ‘stool enema’:ab,ti OR ‘stool infusion’:ab,ti OR ‘stool instillation’:ab,ti OR ‘stool microbial transplantation’:ab,ti OR ‘stool transplant’:ab,ti OR ‘stool transplantation’ AND ‘Systematic review’:ab,ti OR ‘meta-analysis’:ab,ti |
| **Database:** Cochrane<br>**Date limits:**<br>‘bacteriotherapy (feces)’:ab,ti OR ‘faecal bacteriotherapy’:ab,ti OR ‘faecal enema’:ab,ti OR ‘faecal infusion’:ab,ti OR ‘faecal matter transplant’:ab,ti OR ‘faecal microbial transplant’:ab,ti OR ‘faecal microbial transplantation’:ab,ti OR ‘faecal microbiome transplant’:ab,ti OR ‘faecal microbiome transplantation’:ab,ti OR ‘faecal microbiota transplant’:ab,ti OR ‘faecal microbiota transplantation’:ab,ti OR ‘faecal transplant’:ab,ti OR |

‘faecal transplantation’:ab,ti OR ‘fecal bacterial transplant’:ab,ti OR ‘fecal bacterial transplantation’:ab,ti OR ‘fecal bacteriotherapy’:ab,ti OR ‘fecal enema’:ab,ti OR ‘fecal infusion’:ab,ti OR ‘fecal instillation’:ab,ti OR ‘fecal matter transplant’:ab,ti OR ‘fecal matter transplantation’:ab,ti OR ‘fecal microbe transplant’:ab,ti OR ‘fecal microbial transplant’:ab,ti OR ‘fecal microbial transplantation’:ab,ti OR ‘fecal microbiome transplant’:ab,ti OR ‘fecal microbiome transplantation’:ab,ti OR ‘fecal microbiota transplant’:ab,ti OR ‘fecal microbiotal transplant’:ab,ti OR ‘fecal microflora transplantation’:ab,ti OR ‘fecal transfusion’:ab,ti OR ‘fecal transplant’:ab,ti OR ‘fecal transplantation’:ab,ti OR ‘feces bacteriotherapy’:ab,ti OR ‘feces microbe transplantation’:ab,ti OR ‘feces microbiota transplantation’:ab,ti OR ‘feces microflora transplant’:ab,ti OR ‘feces microflora transplantation’:ab,ti OR ‘FMT (fecal microbiota transplantation)’:ab,ti OR ‘gut microbial transplant’:ab,ti OR ‘gut microbial transplantation’:ab,ti OR ‘gut microbiome transplant’:ab,ti OR ‘gut microbiome transplantation’:ab,ti OR ‘gut microbiota transplant’:ab,ti OR ‘gut microbiota transplantation’:ab,ti OR ‘gut microflora transplantation’:ab,ti OR ‘IMT (intestinal microbiota transplantation)’:ab,ti OR ‘intestinal microbe transplantation’:ab,ti OR ‘intestinal microbiota transplant’:ab,ti OR ‘intestinal microbiota transplantation’:ab,ti OR ‘intestinal microflora transplantation’:ab,ti OR ‘rectal bacteriotherapy’:ab,ti OR ‘stool enema’:ab,ti OR ‘stool infusion’:ab,ti OR ‘stool instillation’:ab,ti OR ‘stool microbial transplantation’:ab,ti OR ‘stool transplant’:ab,ti OR ‘stool transplantation’ AND ‘Systematic review’:ab,ti OR ‘meta-analysis’:ab,ti

| Table 2. AMSTAR 2 evaluation method. | |
|---|---|
| Grade | Standard |
| High | No or one non-critical weakness: the systematic review provides an accurate and comprehensive summary of the results of the available studies that address the question of interest. |
| Moderate | More than one non-critical weakness: the systematic review has more than one weakness but no critical flaws. It may provide an accurate summary of the results of the available studies that were included in the review. |
| Low | One critical flaw with or without non-critical weaknesses: the review has a critical flaw and may not provide an accurate and comprehensive summary of the available studies that address the question of interest. |
| Critically low | More than one critical flaw with or without non-critical weaknesses: the review has more than one critical flaw and should not be relied on to provide an accurate and comprehensive summary of the available studies. |

| Table 3. Excluded articles at full-text assessment (n=84) | |
|---|---|
| **First author** | **Reason of exclusion** |
| Madsen[1] | Non-meta-analysis |
| Tan[2] | Non-meta-analysis |
| Fehily[3] | Non-meta-analysis |
| Pession[4] | Non-meta-analysis |
| Leung[5] | Non-meta-analysis |
| Hammeken[6] | Non-meta-analysis |
| Shivaji[7] | Non-meta-analysis |
| Guilfoyle[8] | Non-meta-analysis |
| Marcella[9] | Non-meta-analysis |
| Martínez-González[10] | Non-meta-analysis |
| Chinna[11] | Non-meta-analysis |
| Yang[12] | Non-meta-analysis |
| Stalder[13] | Non-meta-analysis |
| Cold[14] | Non-meta-analysis |
| Hoilat[15] | Non-meta-analysis |
| Feuerstadt[16] | Non-meta-analysis |
| Kayal[17] | Non-meta-analysis |
| Zhang[18] | Non-meta-analysis |
| Du[19] | Non-meta-analysis |
| Pierrard[20] | Non-meta-analysis |
| Hammeken[21] | Non-meta-analysis |
| Saha[22] | Non-meta-analysis |
| Wright[23] | Non-meta-analysis |
| Iqbal[24] | Non-meta-analysis |
| Carrera-Quintanar[25] | Non-meta-analysis |
| Shogbesan[26] | Non-meta-analysis |
| Bafeta[27] | Non-meta-analysis |
| Wang[28] | Non-meta-analysis |
| Chapman[29] | Non-meta-analysis |
| Shogbesan[30] | Non-meta-analysis |
| Drekonja[31] | Non-meta-analysis |
| Rossen[32] | Non-meta-analysis |
| Wang[33] | Non-meta-analysis |
| Cammarota[34] | Non-meta-analysis |
| Drekonja[35] | Non-meta-analysis |
| Sha[36] | Non-meta-analysis |
| Guo[37] | Non-meta-analysis |
| Anderson[38] | Non-meta-analysis |
| **Recurrent Clostridioides difficile infection** | |
| Ramai[39] | Comparison of various methods of faecal microbiota transplantation |
| Tixier[40] | Only one randomized controlled trial |
| Du[41] | No control group (meta-analysis of proportions) |
| Tariq[42] | No outcome of interest |
| Pomares[43] | Comparison of various methods of faecal microbiota transplantation |
| Baunwall[44] | Not the maximum sample size |
| Tariq[45] | No control group (meta-analysis of proportions) |
| Tariq[46] | Abstract only |
| Hong[47] | Abstract only |
| Khan[48] | Comparison of various methods of Faecal microbiota transplantation |
| Tang[49] | Comparison of various methods of Faecal microbiota transplantation |
| Huq[50] | Abstract only |
| Quraishi[51] | Not the maximum sample size |
| Moayyedi[52] | Not the maximum sample size |
| Li[53] | No control group (meta-analysis of proportions) |
| Li[54] | No control group (meta-analysis of proportions) |
| Dakhoul[55] | No control group (meta-analysis of proportions) |
| Dodin[56] | Non-meta-analysis |
| Kassam[57] | No control group (meta-analysis of proportions) |
| Sofi[58] | No outcome of interest (follow-up after faecal microbiota transplantation) |
| Gough[59] | Case series |
| Sofi[60] | Case report |
| **Crohn's disease** | |

| | |
|---|---|
| Cheng[61] | No control group (meta-analysis of proportions) |
| **Antibiotic-resistant bacteria** | |
| Yoon[62] | No control group (meta-analysis of proportions) |
| Tavoukjian[63] | Case series |
| **Ulcerative colitis** | |
| Liu[64] | Not the maximum sample size |
| Dang[65] | Not the maximum sample size |
| Zhao[66] | Not the maximum sample size |
| Lam[67] | Not the maximum sample size |
| Feng[68] | Abstract only |
| Cao[69] | No control group (meta-analysis of proportions) |
| Narula[70] | Not the maximum sample size |
| Keshteli[71] | Letter |
| Costello[72] | Not the maximum sample size |
| Shi[73] | No control group (meta-analysis of proportions) |
| Sun[74] | No control group (meta-analysis of proportions) |
| Scaldaferri[75] | No outcome of interest |
| Green[76] | Not the maximum sample size |
| **Inflammatory bowel disease** | |
| Mocanu[77] | No control group (meta-analysis of proportions) |
| Imdad[78] | Not the maximum sample size |
| Chen[79] | No control group (meta-analysis of proportions) |
| Qazi[80] | No control group (meta-analysis of proportions) |
| Paramsothy[81] | Not the maximum sample size |
| Colman[82] | No control group (meta-analysis of proportions) |
| **Irritable Bowel Syndrome** | |
| Myneedu[83] | Not the maximum sample size |
| Xu[84] | Not the maximum sample size |

**Table 4. The general characteristics of the included meta-analysis.**

| **Source** | **Outcome** | **Year** | **Comparison** | **Number of primary studies** | **Number of cases** | **Follow-up time (wks)** | **Outcome** | **Rude pooled effect estimates** | ***P*-value** | **$I^2$(%)** |
|---|---|---|---|---|---|---|---|---|---|---|
| Dharmaratne[85] | Antibiotic resistance burden | 2021 | FMT VS placebo | 2 | 59 | 28 | Clinical remission | RR = 4.90; 95% CI (1.92-12.50) | 0.0003 | 0 |
| Fang[86] | Functional constipation | 2021 | FMT+ laxative vs laxative | 2 | 163 | 4-12 | Total effective rate | RR=1.35, 95% CI (1.14, 1.60) | 0.0004 | 13 |
| | | | | 3 | 206 | | BFSF score | MD=1.04, 95% CI (0.57, 1.51) | <0.0001 | 76 |
| | | | | 2 | 146 | | Wexner score | MD=-3.25, 95% CI (-5.58, -0.92) | 0.006 | 92 |
| | | | | 2 | 160 | | KESS score | MD=-5.65, 95% CI (-7.62, -3.69) | <0.0001 | 0 |
| | | | | 3 | 246 | | PAC-QOL score | MD=-18.56, 95% CI (-24.63, -10.68) | <0.0001 | 78 |
| | | | | NA | NA | | Adverse effects | NA | NA | NA |
| Caldeira[87] | Inflammatory bowel disease | 2019 | FMT vs placebo | 6 | 355 | 7-12 | Clinical remission | RR=1.70, 95% CI (1.12,2.56) | 0.029 | 45 |
| | | | | 6 | 355 | | Clinical response | RR=1.68, 95% CI (1.04, 2.72) | 0.042 | 55 |
| | | | | NA | NA | | Adverse event | NA | | |
| Tang[88] | Ulcerative colitis | 2020 | FMT vs placebo or vancomycin | 7 | 431 | 4-48 | Clinical remission | RR = 2.29, 95% CI (1.48–3.53) | 0.0002 | 30 |
| | | | | 6 | 416 | | Adverse events | RR = 1.37, 95% CI (0.63–2.96) | 0.43 | 0 |
| | | | | 4 | 267 | | Multi-donor | RR = 1.99, 95% CI (1.17–3.39) | 0.01 | 32 |
| | | | | 2 | 76 | | Single-donor | RR = 1.30, 95% CI (0.98–1.73) | 0.07 | 32 |
| | | | | 5 | 368 | | Lower digestive tract | RR = 1.68, 95% CI (1.09–2.59) | 0.02 | 65 |
| | | | | 2 | 63 | | Up digestive tract | RR = 0.99, 95% CI (0.47–2.09) | 0.97 | 0 |
| Hui[89] | Clostridium difficile infection | 2019 | FMT vs placebo or vancomycin | 8 | 537 | 8-24 | Clinical remission | RR =1.25, 95%CI, 1.01–1.54 | 0.02 | 85 |
| | | | | NA | NA | | Adverse events | NA | | |
| Ianiro[90] | Irritable bowel syndrome | 2019 | FMT vs placebo | 8 | 471 | 8-48 | Clinical remission | RR= 1.08, 95% CI (0.65, 1.77) | 0.65 | 84 |
| | | | | 3 | | | Adverse events | RR=0.93, 95% CI (0.45-1.92) | 0.84 | 61 |

| | | | | | | | | | | |
|---|---|---|---|---|---|---|---|---|---|---|
| | | | | 2 | 100 | | FMT via oral capsules | RR=1.96, 95% CI (1.19-3.20) | 0.008 | 14 |
| | | | | 2 | 103 | | FMT via colonoscopy | RR=0.63, 95% CI (0.43-0.93) | 0.02 | 0 |
| | | | | 1 | 64 | | FMT via nasojejunal tube | RR=0.69, 95% CI (0.46-1.02) | 0.06 | NA |
| Proença[91] | Metabolic Syndrome | 2020 | FMT vs placebo | 6 | 147 | 2-12 | HbA1c | (MD = -1,69, 95% CI (-2.81, -0.56) | 0.003 | 46 |
| | | | | 6 | 146 | | HDL cholesterol | (MD = 0,09, 95% CI (0.02, 0.15) | 0.008 | 5 |
| | | | | 6 | 146 | | LDL cholesterol | (MD = 0.19, 95% CI (0,05, 0.34) | 0.008 | 0 |
| | | | | 6 | 144 | | Fasting glucose | (MD = -0.09, 95% CI (-0,22, 0.04) | 0.16 | 19 |
| | | | | 6 | 119 | | Triglycerides | (MD = -0.02, 95% CI (-0,52, 0.49) | 0.95 | 91 |
| | | | | 5 | 124 | | Total cholesterol | (MD = -0.00, 95% CI (-0,17, 0.16) | 0.97 | 0 |
| | | | | 3 | 78 | | BMI | (MD = -0.85, 95% CI (-2.90, 1.20) | 0.42 | 70 |
| | | | | 4 | 100 | | Weight | (MD = 1.67, 95% CI (-9.50, 12.84) | 0.77 | 86 |
| | | | | 4 | 105 | | HOMA-IR | (MD = -0.35, 95% CI (-1.56, 0.86) | 0.57 | 69 |
| | | | | NA | NA | | Adverse events | NA | NA | NA |

Abbreviations: BFSF, bristol stool form scale; BMI, body mass index; FMT, fecal microbiota transplantation; HbA1c, Hemoglobin A1c; HDL, high density lipoprotein; HOMA-IR, homeostatic model assessment of insulin resistance; KESS, knowles eccersley scott symptom; LDL, low density lipoprotein; MD, mean difference; NA, not applicable; PAC-QOL, patient assessment of constipation quality of life uestionnaire; RR, risk ratio; 95%CI, 95% confidence intervals.

**Table 5. The methodological quality of included meta-analysis using AMSTAR-2**

| References | Item 1 | Item 2 | Item 3 | Item 4 | Item 5 | Item 6 | Item 7 | Item 8 | Item 9 | Item 10 | Item 11 | Item 12 | Item 13 | Item 14 | Item 15 | Item 16 | Overall quality |
|---|---|---|---|---|---|---|---|---|---|---|---|---|---|---|---|---|---|
| Tang[88] | Y | Y | Y | PY | Y | Y | N | Y | Y | N | Y | Y | Y | N | N | Y | Critically low |
| Dharmaratne [85] | Y | Y | Y | PY | Y | Y | N | Y | N | N | Y | N | N | Y | N | Y | Critically low |
| Ianiro[90] | Y | Y | Y | Y | Y | Y | N | Y | Y | N | Y | N | N | Y | N | Y | Critically low |
| Proença[91] | Y | Y | Y | PY | N | Y | N | Y | Y | N | Y | N | N | Y | N | Y | Critically low |
| Hui[89] | Y | Y | Y | PY | N | Y | N | Y | Y | N | Y | Y | Y | Y | Y | Y | Low |
| Fang[86] | Y | Y | Y | PY | Y | Y | N | Y | Y | N | Y | N | N | N | N | Y | Critically low |
| Caldeira[87] | Y | Y | Y | PY | Y | Y | PY | Y | Y | N | Y | N | N | Y | N | Y | Low |

Y: yes; N: no; PY: partial yes

Table 6 GRADE evidence for the included meta-analysis.

| Outcomes | Number of Primary studies | Number of cases | Study design | Risk of bias | Inconsistency | Indirectness | Imprecision | Other considerations | Summary relative risk (95%CI) | Certainty | Importance |
|---|---|---|---|---|---|---|---|---|---|---|---|
| Antibiotic-resistant bacteria | 2 | 69 | randomised trials | very serious[1] | not serious | not serious | serious[3] | none | 5.67 (2.20, 14.63) | ⊕⊝⊝⊝ very low | 7 |
| Irritable Bowel Syndrome | 8 | 471 | randomised trials | serious[2] | very serious[4] | not serious | not serious | none | 1.08 (0.65, 1.77) | ⊕⊝⊝⊝ very low | 7 |
| Clostridioides difficile infection | 8 | 452 | randomised trials | serious[2] | not serious | not serious | not serious | none | 1.74 (1.37, 2.22) | ⊕⊕⊕⊝ moderate | 8 |
| functional constipation | 5 | 128 | randomised trials | very serious[1] | not serious | not serious | serious[3] | none | 1.35 (1.14, 1.60) | ⊕⊝⊝⊝ very low | 8 |
| Ulcerative colitis | 14 | 675 | randomised trials | serious[2] | not serious | not serious | not serious | none | 2.05 (1.05, 3.05) | ⊕⊕⊕⊝ moderate | 8 |
| Inflammatory bowel disease | 6 | 355 | randomised trials | serious[2] | not serious | not serious | not serious | none | 1.70 (1.12, 2.56) | ⊕⊕⊕⊝ moderate | 8 |

Explanation.
[1]Poor information regarding randomization,allocation concealment and blinding.
[2]Poor information regarding randomization and allocation concealment.
[3]Small sample size (two arm with less than 100 participants).
[4]$I^2$ >75 %.

[66] Zhao HL,Chen SZ,Xu HM,Zhou YL,He J,Huang HL,Xu J,Nie YQ. Efficacy and safety of fecal microbiota transplantation for treating patients with ulcerative colitis: A systematic review and meta-analysis.J Dig

Dis.2020; 21,534-548.

[67] Lam WC,Zhao C,Ma WJ,Yao L. The Clinical and Steroid-Free Remission of Fecal Microbiota Transplantation to Patients with Ulcerative Colitis: A Meta-Analysis.Gastroenterol Res Pract.2019; 2019,1287493.

[68] Feng X,Li S. Fecal microbial transplantation for the treatment of ulcerative colitis :a syetematic review and meta analysis.J. Gastroen. Hepatol.2019; 34,135.

[69] Cao Y,Zhang B,Wu Y,Wang Q,Wang J,Shen F. The Value of Fecal Microbiota Transplantation in the Treatment of Ulcerative Colitis Patients: A Systematic Review and Meta-Analysis.Gastroenterol Res Pract.2018; 2018,5480961.

[70] Narula N,Kassam Z,Yuan Y,Colombel JF,Ponsioen C,Reinisch W,Moayyedi P. Systematic Review and Meta-analysis: Fecal Microbiota Transplantation for Treatment of Active Ulcerative Colitis.Inflamm. Bowel Dis.2017; 23,1702-1709.

[71] Keshteli AH,Millan B,Madsen KL. Pretreatment with antibiotics may enhance the efficacy of fecal microbiota transplantation in ulcerative colitis: a meta-analysis.Mucosal Immunol.2017; 10,565-566.

[72] Costello SP,Soo W,Bryant RV,Jairath V,Hart AL,Andrews JM. Systematic review with meta-analysis: faecal microbiota transplantation for the induction of remission for active ulcerative colitis.Alimentary Pharmacology and Therapeutics.2017; 46,213-224.

[73] Shi Y,Dong Y,Huang W,Zhu D,Mao H,Su P. Fecal Microbiota Transplantation for Ulcerative Colitis: A Systematic Review and Meta-Analysis.PLoS One.2016; 11,e157259.

[74] Sun D,Li W,Li S,Cen Y,Xu Q,Li Y,Sun Y,Qi Y,Lin Y,Yang T,Xu P,Lu Q. Fecal Microbiota Transplantation as a Novel Therapy for Ulcerative Colitis: A Systematic Review and Meta-Analysis.Medicine (Baltimore).2016; 95,e3765.

[75] Scaldaferri F,Pecere S,Petito V,Zambrano D,Fiore L,Lopetuso LR,Schiavoni E,Bruno G,Gerardi V,Laterza L,Pizzoferrato M,Ianiro G,Stojanovic J,Poscia A,Papa A,Paroni SF,Sanguinetti M,Masucci L,Cammarota G,Gasbarrini A. Efficacy and Mechanisms of Action of Fecal Microbiota Transplantation in Ulcerative Colitis: Pitfalls and Promises From a First Meta-Analysis.Transplant Proc.2016; 48,402-407.

[76] Green JE,Davis JA,Berk M,Hair C,Loughman A,Castle D,Athan E,Nierenberg AA,Cryan JF,Jacka F,Marx W. Efficacy and safety of fecal microbiota transplantation for the treatment of diseases other than Clostridium difficile infection: a systematic review and meta-analysis.Gut Microbes.2020; 12,1-25.

[77] Mocanu V,Rajaruban S,Dang J,Kung JY,Deehan EC,Madsen KL. Repeated Fecal Microbial Transplantations and Antibiotic Pre-Treatment Are Linked to Improved Clinical Response and Remission in Inflammatory Bowel Disease: A Systematic Review and Pooled Proportion Meta-Analysis.J Clin Med.2021; 10.

[78] Imdad A,Nicholson MR,Tanner-Smith EE,Zackular JP,Gomez-Duarte OG,Beaulieu DB,Acra S. Fecal transplantation for treatment of inflammatory bowel disease.Cochrane Database Syst Rev.2018; 11,D12774.

[79] Chen T,Zhou Q,Zhang D,Jiang F,Wu J,Zhou JY,Zheng X,Chen YG. Effect of faecal microbiota transplantation for treatment of Clostridium difficile infection in patients with inflammatory bowel disease: A systematic review and meta-analysis of cohort studies.Journal of Crohn's and Colitis.2018; 12,710-717.

[80] Qazi T,Amaratunga T,Barnes EL,Fischer M,Kassam Z,Allegretti JR. The risk of inflammatory bowel disease flares after fecal microbiota transplantation: Systematic review and meta-analysis.Gut Microbes.2017; 8,574-588.

[81] Paramsothy S,Paramsothy R,Rubin DT,Kamm MA,Kaakoush NO,Mitchell HM,Castaño-Rodríguez N.

Faecal microbiota transplantation for inflammatory bowel disease: A systematic review and meta-analysis.Journal of Crohn's and Colitis.2017; 11,1180-1199.
[82] Colman RJ,Rubin DT. Fecal microbiota transplantation as therapy for inflammatory bowel disease: a systematic review and meta-analysis.J. Crohns Colitis.2014; 8,1569-1581.
[83] Myneedu K,Deoker A,Schmulson MJ,Bashashati M. Fecal microbiota transplantation in irritable bowel syndrome: A systematic review and meta-analysis.United European Gastroenterol J.2019; 7,1033-1041.
[84] Xu D,Chen VL,Steiner CA,Berinstein JA,Eswaran S,Waljee AK,Higgins P,Owyang C. Efficacy of Fecal Microbiota Transplantation in Irritable Bowel Syndrome: A Systematic Review and Meta-Analysis.Am. J. Gastroenterol.2019; 114,1043-1050.
[85] Dharmaratne P, Rahman N, Leung A, Ip M. Is there a role of faecal microbiota transplantation in reducing antibiotic resistance burden in gut? A systematic review and Meta-analysis. Annals of medicine (Helsinki). 2021; 53: 662-81.
[86] Fang S, Wu S, Ji L, Fan Y, Wang X, Yang K. The combined therapy of fecal microbiota transplantation and laxatives for functional constipation in adults. Medicine. 2021; 100: e25390.
[87] Caldeira LDF, Borba HH, Tonin FS, Wiens A, Fernandez-Llimos F, Pontarolo R. Fecal microbiota transplantation in inflammatory bowel disease patients: A systematic review and meta-analysis. Plos One. 2020; 15: e238910.
[88] Tang L, Feng W, Cheng J, Gong Y. Clinical remission of ulcerative colitis after different modes of faecal microbiota transplantation: A meta-analysis. Int J Colorectal Dis. 2020; 35: 1025-34.
[89] Hui W, Li T, Liu W, Zhou C, Gao F. Fecal microbiota transplantation for treatment of recurrent C. Difficile infection: An updated randomized controlled trial meta-analysis. Plos One. 2019; 14: e210016.
[90] Ianiro G, Eusebi LH, Black CJ, Gasbarrini A, Cammarota G, Ford AC. Systematic review with meta-analysis: Efficacy of faecal microbiota transplantation for the treatment of irritable bowel syndrome. Aliment Pharm Ther. 2019; 50: 240-8.
[91] Proença IM, Allegretti JR, Bernardo WM, de Moura DTH, Ponte Neto AM, Matsubayashi CO, et al. Fecal microbiota transplantation improves metabolic syndrome parameters: Systematic review with meta-analysis based on randomized clinical trials. Nutrition research.2020; 83: 1-14.
